# Exploring the utility and scalability of entrustable professional activities beyond medical education: A scoping review protocol

**DOI:** 10.1101/2025.08.10.25333060

**Authors:** Anjali Jagannathan, Julia Micallef, Otto Sanchez, Adam Dubrowski

**Author notes:** Corresponding Author (AJ). These authors contributed equally to this work (Anjali Jagannathan, Otto Sanchez, Adam Dubrowski).

## Abstract

Competency-based education (CBE) is gaining prominence in post-secondary education due to its effectiveness in optimizing workforce readiness. However, assessing competence and workforce readiness presents a significant challenge. Entrustable professional activities (EPAs) offer an effective solution by translating competencies into observable, measurable, and entrustable tasks. While EPAs have been widely adopted in medical education, their application in non-clinical educational contexts remains minimal. This scoping review aims to map the diffusion of EPAs beyond medicine and assess their scalability to undergraduate non-clinical minor pathways, which are ideal piloting sites for EPA-based assessment frameworks. Guided by Rogers’ Diffusion of Innovations theory, this review will identify the barriers and facilitators influencing the adoption of EPAs in non-clinical educational contexts. Following Arksey and O’Malley’s framework, this review will progress through five stages: identifying the research question, identifying relevant studies, study selection, charting the data, and reporting results. Searches will be conducted across Ovid MEDLINE, CINAHL, Scopus, ERIC, and grey literature. The research question is: “How have EPAs been developed and implemented outside of medical education, and what factors influence their adoption in non-clinical training programs?” The search will include articles describing the development or implementation of EPAs outside medicine and will exclude irrelevant studies. Titles and abstracts will be reviewed first, followed by a full-text examination of relevant articles. Data will be extracted, organized, and summarized to highlight key findings. Two reviewers will ensure quality during screening and data extraction. Findings from this scoping review will inform the rationale for scaling EPAs to non-clinical training programs and offer insights to guide the adaptation, development, and implementation of EPA-based assessment frameworks in undergraduate non-clinical minor pathways, addressing pressing challenges in CBE and creating a replicable model for future program development. Findings from this scoping review will be submitted for publication in a scientific journal.

## Introduction

Competency-based education (CBE) is gaining prominence in post-secondary education due to its emphasis on demonstrated competence, with increasing empirical support for its effectiveness in optimizing workforce readiness [1]. However, while competency frameworks outline the knowledge, skills, and attitudes (KSAs) essential for professional practice, KSAs remain intrinsic attributes that cannot be directly observed in real-world performance [2]. Hence, conducting reliable, defensible assessments of competence and workforce readiness remains a persistent challenge undermining CBE [2]. In medical education, Ten Cate [3] addressed this challenge by introducing Entrustable Professional Activities (EPAs), which operationalize competencies as observable, measurable, and entrustable tasks. The strength of EPAs lies in their ability to shift the focus of assessment from isolated KSAs to the real-world execution of professional activities, bridging the gap between competency frameworks and professional practice [2]. In medical education, EPA proficiency is developed longitudinally across curricula aligned with Miller’s framework [4], where learners progress from classroom-based education, to simulation training, to real-world performance [2], with assessments embedded at each stage. This structure ensures learners demonstrate both canonical competence (the KSAs required to perform EPAs) and contextual competence (the ability to apply integrated KSAs to dynamic real-world practice) [5]. Within this structure, EPAs enable entrustment-based assessment, in which trainees are progressively granted autonomy as they develop proficiency and are entrusted to perform an EPA unsupervised only once competence has been demonstrated [2], ensuring workforce readiness upon graduation.

Despite their demonstrated efficacy and widespread implementation in medical education [6], EPAs remain largely confined to clinical training programs that lead to licensure in medicine, nursing, or allied health professions (i.e., dietetics, physiotherapy, pharmacy etc.).

These programs are typically accredited and include formal clinical placements as part of their curriculum. The adoption of EPAs beyond medicine in non-clinical domains, such as teaching in the health professions and undergraduate degrees in health sciences, biological sciences, and arts, has been minimal. This limited uptake reflects both a lack of established frameworks for translating EPAs into non-clinical domains and an underexplored opportunity to use EPAs to bridge competencies and professional practice in diverse educational settings. As EPAs continue to evolve as an effective assessment tool, expanding their use beyond clinical professions represents a critical next step for educational innovation and equity across disciplines. While previous scoping reviews have explored the use of EPAs across clinical disciplines [7,8], to our knowledge, no review has mapped their application or assessed their scalability to non-clinical contexts. EPAs offer a promising solution to assessment challenges in CBE across many disciplines, as the underlying logic of entrustment is broadly applicable to any professional field where real-world performance carries risk. This includes a broad range of non-clinical programs situated within university-based pathways where technical, pedagogical, and organizational competencies must be demonstrated with confidence and accountability. Errors in these contexts may not directly harm patients, as is the case in clinical training, but can undermine system integrity, educational outcomes, or public trust. As such, EPA-based assessment frameworks present a compelling opportunity for adaptation beyond clinical training contexts. Minor pathways embedded within larger undergraduate degrees, such as minors in recreation therapy, public health, and simulation operations, offer ideal pilot sites for this innovation, as these sub-programs frequently grow into full professional programs over time. Introducing EPAs within minor pathways will allow for effective competency assessment and workforce alignment from the outset, ensuring that when they transition to professional program status, graduates are prepared for professional practice. By developing and implementing EPAs in these non-clinical minor pathways, institutions can address long-standing challenges in CBE while building scalable, transferable models to guide future program development. However, successful implementation will require careful contextual adaptation, accounting for institutional policy environments, supervision and autonomy structures, and potential resistance from involved parties. This underscores the importance of examining the barriers and facilitators that influence the adoption and diffusion of EPAs across non-clinical educational contexts.

To guide this inquiry, this scoping review will be informed by Rogers’ Diffusion of Innovations (DoI) theory [9], a conceptual framework for understanding how innovations spread through systems over time. Rogers identified five core factors that enable or constrain the adoption of innovations: (1) Relative advantage, the perceived benefits over existing approaches; Compatibility, the alignment with needs and values; (3) Complexity, the perceived ease of understanding, utilization, and implementation; (4) Trialability, the degree to which the innovation can be tested and piloted; and (5) Observability, the visibility of the benefits and results. Given the limited literature on EPAs outside medical education, a scoping review is the optimal methodology to explore their adoption and scalability beyond this context [10]. By applying DoI theory, this review will not only map the diffusion of EPAs beyond medical education but also assess the contextual factors that affect their uptake, success, and adaptation to novel contexts, highlighting lessons learned from early adopters and uncovering actionable strategies for successful development and implementation. If the review confirms the scarcity of existing literature on and evidence of scalability of EPAs to non-clinical educational contexts, it will inform the rationale for scaling, developing and implementing EPA-based assessment frameworks for non-clinical training programs, with undergraduate minor pathways providing the optimal piloting site for future program development. The research question explored through this scoping review will be: “*How have EPAs been developed and implemented outside of medical education, and what contextual factors influence their adoption in non-clinical training programs?”*

### Protocol Design

This scoping review will be guided by the methodological framework proposed by Arksey and O’Malley [11], which outlines a five-stage process: 1) identifying the research question, 2) identifying relevant studies, 3) selecting studies for inclusion, 4) charting the data, and 5) collating, summarizing, and reporting the results. To ensure methodological quality, we will also draw upon the JBI Manual for Evidence Synthesis [12] and report our results in accordance with the PRISMA-ScR (Preferred Reporting Items for Systematic Reviews and Meta-Analyses – Scoping Review) Checklist [13]. Given that the review will exclusively rely on analysis of publicly available materials, no ethics approval is required. Additionally, a health science librarian will be consulted to support database selection and search strategy development. As of July 2025, stage 1 has been completed, having identified the research question, and stage 2 is in progress, having begun initial consultations with a health science librarian to develop a search strategy and select databases to identify relevant studies. The following subsections describe each stage in detail, outlining the rationale, scope, and intended outputs.

### Stage 1: Identifying the research question

Guided by Rogers’ DoI theory [9], the purpose of this scoping review is to explore the existing literature to identify how and where EPAs are used beyond medical education, and examine contextual factors that influence successful adaptation, uptake, and implementation in non-clinical training contexts. In consultation with the research team, the following research question has been developed: “*How have EPAs been developed and implemented outside of medical education, and what contextual factors influence their adoption in non-clinical training programs*?” To address this question, the objectives of the proposed scoping review are 1) to map the diffusion of EPAs beyond medical education, 2) to determine how EPAs have been developed and implemented across curricula outside medical education, and 3) to explore evidence of scalability to non-clinical educational contexts. The findings of this review will be interpreted through the lens of five core attributes identified by Rogers. If the review confirms the scarcity of EPAs in non-clinical training and evidence of scalability, it will inform the rationale and provide insights to guide their successful adaptation, development, and implementation in these contexts.

### Stage 2: Identifying relevant studies

Four literature databases covering the medical, nursing, science and technology, and education fields will be searched for this scoping review: Ovid MEDLINE, CINAHL, Scopus, and ERIC. Concepts derived from the research question will be used to develop free-text codes for the search strategy on Ovid MEDLINE, which will then be translated for the other three remaining databases. The search strategy will include keywords such as “entrustable professional activities”, “entrustment”, “health professions education”, “health sciences education”, “non-clinical program”, “educator”, “facilitator” and their synonyms, and a search filter developed at the University of Alberta will be modified and used to derive studies related to health students, using terms such as “nursing”, “pharmacy”, “audiology”, “premedical”, “public health”, “residency”, internship”, “trainee”, and their synonyms [14]. These free-text codes will be combined with database-specific controlled subject headings to formulate the search strategy on each of the four published literature databases. The detailed search strategy for Ovid MEDLINE is presented in the S1 Table. To mitigate the possibility of limited results from published literature, the following grey literature databases will be searched: Grey Matters (CADTH), OpenGrey, and Google Scholar. This search strategy was developed in consultation with a health science librarian in accordance with the Peer Review of Electronic Search Strategies (PRESS) guidelines [15]. No time limits will be applied to the search strategy. While EPAs were formally introduced by Ten Cate in 2005, this review seeks to identify not only how and where EPAs have been developed and implemented beyond medicine but also the contextual factors that influence their adoption. Removing temporal restrictions allows for the identification of conceptual antecedents that may align with the principles of EPAs and highlight lessons learned from early efforts to develop and implement similar entrustment-based assessment frameworks, thereby providing a more comprehensive understanding of the barriers and facilitators influencing EPA adoption across non-clinical training programs. Furthermore, the reference lists of relevant articles will be manually searched to identify any new articles that address the research question.

### Stage 3: Study selection

The citation management software EndNote 21[16] will be used for the initial screening to organize the selected articles and remove duplicates. The results will be imported into EPPI-Reviewer software [17] for further screening and selection. The screening of the articles will follow a two-step process. Initially, the titles and abstracts will be reviewed to determine the eligibility of each article, and those that do not meet the inclusion criteria will be excluded. Secondly, a full-text screening of the articles that pass the initial screening will be reviewed in full-text. Only relevant articles identified in step two will be included in the review. The study selection process will be reported using the PRISMA flow diagram. Articles will be selected based on specific inclusion and exclusion criteria, ensuring they address the research question. The inclusion criteria will include a need for the article to describe the development or implementation of EPAs in a training context outside medical education. Exclusion criteria will include editorial articles and conference abstracts and posters. Study designs that will be accepted are surveys, case studies, systematic reviews, scoping reviews, mixed methods studies, and commentaries/program evaluations.

### Stage 4: Charting the data

The data will be extracted and charted using the EPPI-Reviewing software [17]. The following key elements will be extracted from the articles: author details, country of origin, study objective/purpose, study design, the context in which EPAs are being used; the strategies employed for their development and implementation across curricula; the evidence of their scalability to non-clinical training contexts categorized by the five attributes of DoI identified by Rogers, and any other key findings related to the research question.

### Stage 5: Collating, summarizing, and reporting the results

In accordance with the purpose of this scoping review, the context in which EPAs are being used, the strategies used for development and implementation across curricula, and evidence of EPA scalability to non-clinical training contexts according to the five attributes of DoI identified by Rogers, will be reported. Quantitative and qualitative data will be summarized descriptively in text, and presented in tables and graphs where appropriate, adhering to the Joanna Briggs Institute Manual for Evidence Synthesis and the PRISMA-ScR Checklist [13].

### Quality assurance

The citation management tool, EndNote 21 [16] will be used to organize articles after executing the search strategy, to create references for the selected articles. The EPPI-Reviewer software [17], a web-based program for managing and analyzing data in literature reviews, will be used to screen and select articles. Two reviewers (AJ & JM) will independently screen the titles and abstracts of a subset of the search results (10%) to assess eligibility criteria and perform a quality check. Then, the reviewers will discuss their ratings and refine the eligibility criteria as needed. Should any disagreements arise, they will be resolved by a third reviewer. Following this, one reviewer (AJ) will screen the remaining references, with any questions checked by the second reviewer (JM). During the charting stage, data will be extracted from 100% of the studies by one reviewer (AJ) and from two random samples of 5% each by the second reviewer (JM) to ensure the quality of the charting process.

## Dissemination

By systematically examining existing literature, this scoping review aims to map the diffusion of EPAs beyond medical education, identify how they have been developed and implemented across curricula in these contexts, and explore evidence of scalability to non-clinical programs. Rogers’ DoI theory will serve as an interpretive framework to categorize and analyze patterns within the findings. By assessing the current state of research and identifying gaps, this review will inform the rationale for scaling EPAs to non-clinical educational contexts and provide insights to guide their successful adaptation, development, and implementation in non-clinical undergraduate minor pathways. This review has the potential to inform future research and practice in expanding EPAs further beyond their medical education origins.

Furthermore, it will offer insights to guide the development and implementation of EPA-based assessment frameworks within undergraduate minor pathways, creating a replicable model for future non-clinical program development. The estimated date of the scoping review’s completion is in Spring 2026. The results of this review will be submitted for publication in a peer-reviewed journal, contributing to and guiding future advancements in the broader field of EPAs and competency-based education.

## Strengths and limitations

This scoping review will be the first to examine literature on the development and implementation of EPAs beyond medical origins, inclusive of non-clinical contexts, and assess scalability to these contexts. This scoping review protocol has several strengths. It features a comprehensive search strategy across multiple databases and grey literature sources, which enhances the likelihood of capturing a wide range of relevant studies. The research questions and objectives are clearly defined, providing a focused direction for the scoping review. It utilizes DoI theory, a widely used conceptual framework, to categorize and interpret the findings, adding depth to the analysis. Furthermore, the review protocol adheres to frameworks and guidelines, such as those established by Arksey and O’Malley [11] and the PRISMA-ScR Checklist [13], ensuring a systematic and transparent approach. The two-step screening process involving independent reviews and incorporating quality assurance adds rigor to the study selection.

Additionally, the review’s reliability is further strengthened by the use of EndNote 21 [16] and EPPI-Reviewer software [17]. Despite these strengths, this protocol possesses certain limitations. A preliminary scan shows a paucity of literature on the use of EPAs in non-clinical contexts, which could affect the comprehensiveness of the review. Moreover, the resource-intensive nature of the review process, involving multiple stages and reviewers, could pose challenges in terms of time and resource management.

## Supporting information

https://www2.cloud.editorialmanager.com/pone/download.aspx?id=41458541&guid=a47e1c61-62d2-42de-b486-8d9e5648e87b&scheme=1

https://www2.cloud.editorialmanager.com/pone/download.aspx?id=41458558&guid=ff306f7d-aa2f-496b-a835-67de888ef437&scheme=1

## Data Availability

No datasets were generated or analysed during the current study. All relevant data from this study will be made available upon study completion.

## Supporting Information

**S1 Table. Search strategy for Ovid MEDLINE**

**S2 Checklist. PRISMA-P checklist**

## References

1. Frank JR, Snell LS, Ten Cate O, Holmboe ES, Carraccio C, Swing SR. Competency-based medical education: theory to practice. Med Teach. 2010 Jul 27;32(8): 638–45. 10.3109/0142159X.2010.501190

2. Ten Cate O et al., (eds) Entrustable Professional Activities and Entrustment Decision-Making in Health Professions Education. London: Ubiquity Press; 2024. 10.5334/bdc.b

3. Ten Cate O. Entrustability of professional activities and competency-based training. Med Educ. 2005 Nov 25;39(12): 1176–7. 10.1111/j.1365-2929.2005.02341.x

4. Miller GE. The assessment of clinical skills/competence/performance. Acad Med. 1990 Sep;65(9): S63–7. 10.1097/00001888-199009000-00045

5. Teunissen PW, Watling CJ, Schrewe B, Asgarova S, Ellaway R, Myers K, Topps M, Bates J. Contextual competence: how residents develop competent performance in new settings. Med Educ. 2021 Feb 25;55(9): 1100–1109. 10.1111/medu.14517

6. Park S, Chiang NJ, Nyhof-Young J, Rojas D, Lazor, J, Sirianni G. Entrustable professional activities in undergraduate medical education: a needs assessment of medical students and faculty. Educ Health Prof. 2023;6(2): 92–6. 10.4103/EHP.EHP_3_23

7. Bramley AL, McKenna L. Entrustable professional activities in entry-level health professional education: a scoping review. Med Educ. 2021 Apr 21;55(9): 1011–1032. 10.1111/medu.14539

8. Shorey S, Lau TC, Lau ST, Ang E. Entrustable professional activities in health care education: a scoping review. Med Educ, 2019 Apr 4;53(8): 766–777. 10.1111/medu.13879

9. Rogers EM. Diffusion of Innovations, 5th Edition. New York: Free Press; 2023. https://teddykw2.wordpress.com/wp-content/uploads/2012/07/everett-m-rogers-diffusion-of-innovations.pdf

10. Grant MJ, Booth A. A typology of reviews: An analysis of 14 review types and associated methodologies. Health Info Libr J. 2009 May 27;26(2): 91–108. 10.1111/j.1471-1842.2009.00848.x

11. Arksey H, O’Malley L. Scoping studies: Towards a methodological framework. Int J Soc Res Methodol. 2005;8(1): 19–32. 10.1080/1364557032000119616

12. Aromataris E, Munn Z (eds). JBI manual for evidence synthesis. JBI. 2020. Available from: 10.46658/JBIMES-20-01

13. Tricco AC, Lillie E, Zarin W, O’Brien KK, Colquhoun H, Levac D, et al. PRISMA extension for scoping reviews (PRISMA-ScR): checklist and explanation. Ann Intern Med. 2018 Oct 2;169(7):467–473. Available from: pmid:30178033. 10.7326/M18-0850

14. Kung JY. Filter to Retrieve Studies Related to Health Students from the OVID EMBASE Database. Geoffrey & Robyn Sperber Health Sciences Library, University of Alberta. 2023. Available from: https://docs.google.com/document/d/1CzcaRNxmLBT_5vEdED_98BMOUwcAY-Tuzt855hEd7gg/edit#heading=h.ldbxqb34y1kj

15. McGowan J, Sampson M, Salzwedel D, Cogo E, Foerster V, Lefebvre C. PRESS peer review of electronic search strategies: 2015 guideline statement. J Clin Epidemiol. 2016 Jul;75:40–46. 10.1016/j.jclinepi.2016.01.021

16. The EndNote Team. EndNote. Version EndNote 20 [software]. 2023 [cited 2025 July 13]. Available from: https://eppi.ioe.ac.uk/CMS/Default.aspx?alias=eppi.ioe.ac.uk/cms/er4&amp

17. Thomas J, Graziosi S, Brunton J, Ghouze Z, O’Driscoll P, Bond M, et al. EPPI-Reviewer: advanced software for systematic reviews, maps and evidence synthesis. EPPI-Centre. 2022 [cited 2025 July 13]. Available from: https://eppi.ioe.ac.uk/CMS/Default.aspx?alias=eppi.ioe.ac.uk/cms/er4&

