## Supplementary material for "Exploring the utility and scalability of entrustable professional activities beyond medical education: A scoping review protocol": https://www2.cloud.editorialmanager.com/pone/download.aspx?id=41458541&guid=a47e1c61-62d2-42de-b486-8d9e5648e87b&scheme=1

**S1 Table. Search strategy for Ovid MEDLINE**

| Line # | Code |
|---|---|
| 1 | entrustable professional activit*.mp. |
| 2 | entrustment.mp. |
| 3 | 1 or 2 |
| 4 | non-clinical program*.mp. |
| 5 | health professional education*.mp. |
| 6 | health professions education*.mp. |
| 7 | HPE.mp. |
| 8 | health sciences education*.mp. |
| 9 | (facilitator* or director* or teacher* or educator*).mp. |
| 10 | students, Health Occupations / or students, dental/ or students,nursing/ or students, pharmacy/ or students, premedical/ or students, public health/ |
| 11 | ((audiology or chiropract* or dental or dentistry or dietetics or health or health care or healthcare or midwifery or nurs* or optometr* or paramedic*or pharmac* or psycholog* or public health or therapy) adj3 (intern or interns or internship* or learner* or resident* or residency or student* or trainee* or training* or education*)).mp. |
| 12 | 10 or 11 |
| 13 | education, predental/ or education, premedical/ or exp education, dental/ or exp "education, biological and biomedical sciences, graduate"/ or education, dental, graduate/ or education, nursing, graduate/ or education, pharmacy, graduate/ or exp education, nursing/ or exp education, pharmacy/ or education public health professional/ or education, veterinary/ or exp internship, nonmedical |
| 14 | 4 or 5 or 6 or 7 or 8 or 9 or 12 or 13 |
| 15 | 3 and 14 |
